# Traumatic microbleeds are associated with unfavorable outcome out to 1 year after mild traumatic brain injury: A TRACK-TBI Study

**DOI:** 10.64898/2026.11.15.26367999

**Authors:** Esther L. Yuh, Sonia Jain, Xiaoying Sun, Sandra P. Rincon, Pratik Mukherjee, John K. Yue, Amy J. Markowitz, Christine L. Mac Donald, Gabriela G. Satris, Sabrina R. Taylor, Joseph T. Giacino, Michael W. Mccrea, Murray B. Stein, Nancy R. Temkin, Ramon Diaz-Arrastia, Claudia S. Robertson, David O. Okonkwo, Geoffrey T. Manley, The Track-TBI Investigators

## Abstract

**Importance:** Traumatic microbleeds (TMB) are commonly observed in mild TBI (mTBI). However, questions remain regarding their long-term prognostic significance.

**Objective:** Determine whether traumatic microbleeds predict unfavorable outcome after mild traumatic brain injury (mTBI)

**Design:** The prospective longitudinal observational <u>T</u>ransforming <u>R</u>esearch <u>a</u>nd <u>C</u>linical <u>K</u>nowledge in <u>T</u>raumatic <u>B</u>rain Injury (TRACK-TBI) study enrolled acute TBI patients presenting to Emergency Departments (ED) at 18 U.S. Level 1 trauma centers between 2/26/2014-08/08/2018. CT and MRI were evaluated using the National Institutes of Health-National Institute of Neurological Disorders and Stroke (NIH-NINDS) TBI Common Data Elements. Outcome was assessed using the Glasgow Outcome Scale–Extended (GOSE-TBI) at 2 weeks and 3, 6, and 12 months post-injury.

**Setting:** 18 U.S. Level 1 trauma centers

**Participants:** 929 GCS 13-15 patients ≥17 years old (38.5 ± 16.5 years, 66% male) who underwent clinical head CT ≤24 hours and brain MRI at 2 weeks post-injury and had at least one GOSE-TBI outcome score during 1 year follow-up.

**Exposure(s):** Acute non-penetrating head trauma

**Main Outcome(s) and Measure(s):** Frequency and co-occurrence of CT and MR features; incomplete recovery (GOSE-TBI <8 vs. =8) and unfavorable outcome (GOSE-TBI <5 vs. ≥5) at 2 weeks and 3, 6, and 12 months

**Results:** Generalized estimating equation models showed that TMBs and/or IVH were strongly associated with unfavorable outcome (GOSE-TBI <5 vs. ≥5) at 3, 6, and 12 months post-injury, but not with incomplete recovery (GOSE-TBI <8 vs. =8). For prediction of GOSE-TBI <5 vs. ≥5, TMBs surpassed the importance of all demographic and baseline clinical predictors throughout the 1-year follow-up period. For prediction of incomplete recovery (GOSE-TBI <8 vs. =8), brain contusions, subarachnoid hemorrhage, and subdural hematoma were more important throughout the 12-month follow-up period.

**Conclusions and Relevance:** Traumatic microbleeds (TMB) are common in mTBI. The presence of ≥4 TMBs and/or IVH is strongly associated with unfavorable outcome (GOSE-TBI <5) after mTBI out to at least 1 year, even after controlling for other neuroimaging abnormalities and demographic and clinical predictors. Better understanding of the implications of TMBs for long-term functional outcome may inform rehabilitation planning.

**KEY POINTS:** *Question:* Do traumatic microbleeds predict unfavorable outcome after mild traumatic brain injury (mTBI)?

*Findings:* In this cohort study of 929 mTBI patients, traumatic microbleeds were common; ≥4 TMBs in at least 2 distinct regions of the brain were observed in 11.3% of participants on brain MRI performed 2 weeks post-injury. TMBs were associated more strongly with intraventricular hemorrhage (IVH) than with other neuroimaging findings including brain contusions, subarachnoid hemorrhage, and subdural and epidural hematomas. The presence of ≥4 TMBs on brain MRI, or any TMB or IVH on head CT, were strongly associated with unfavorable outcome (GOSE-TBI <5) out to 12 months after mTBI. Specifically, these neuroimaging findings had odds ratios (OR) of 8.3 [2.4-29.2], 7.5 [1.8-30.8], and 5.4 [1.6-18.0] for GOSE-TBI <5 vs. ≥5 at post-injury months 3, 6, and 12, respectively. These far exceeded the ORs for other predictors including other CT and MRI findings, admission GCS score (13 vs15), age, sex, years of education, history of neuropsychiatric disorder, history of prior TBI, and contemporaneous major extracranial injury.

*Meaning:* TMBs and IVH are associated with each other, and with unfavorable outcome (GOSE-TBI <5) out to at least 12 months after mild TBI. The association persists after controlling for other CT and MRI findings, socio-demographic factors, and clinical characteristics.

## INTRODUCTION

It is known that magnetic resonance imaging (MRI) has superior sensitivity over computed tomography (CT) for small cortical contusions and for markers of rotational injury such as traumatic microbleeds (TMB) after acute TBI.^1–3^ However, questions remain regarding the long-term prognostic significance of these MRI findings.

Previous studies of the association between TMBs on MRI and outcome have had conflicting results. These include univariable analyses,^1,4–8^ as well as studies in which MRI findings were included in multivariable models of outcome prediction.^1,9–15^ Some of these studies reported that traumatic lesions on MRI were associated with worse outcome after TBI,^4,10,11,13,15^ but many found no significant association.^1,4–9,12^ One study demonstrated that TMBs, while common in mTBI, had no association with outcome, even at 2 weeks post-injury ^6^ In contrast, another study demonstrated that TMBs on MRI, though more prevalent in severe TBI (GCS 3-8), are common across all GCS scores, and are associated with worse outcome (Glasgow Outcome Scale-Extended (GOSE-TBI ≤6) at intermediate-term follow-up (ranging from 1 to 3 months);^10^ however, longer term outcome data beyond 3 months postinjury were not available in that study.

In this study, we sought to determine the prognostic significance of TMBs for long-term outcome out to 1 year, when added to other CT and MRI findings and socio-demographic and baseline clinical features. We also sought to determine the additive prognostic value of early brain MRI when added to head CT findings.

## PATIENTS AND METHODS

### Study population

#### Transforming Research and Clinical Knowledge in Traumatic Brain Injury (TRACK-TBI)

enrolled TBI participants who presented to the emergency departments (ED) at 1 of 18 U.S. Level 1 trauma centers (**Supplementary Table 1**) and were treated along 1 of 3 care pathways (ED discharge, hospital admission without ICU, hospital admission with ICU). The *inclusion criterion* for TRACK-TBI was presentation to a participating center within 24 hours of injury with clinical indication for head CT under American College of Emergency Medicine/Centers for Disease Control and Prevention guidelines.^16^ *Exclusion criteria* included pregnancy, incarceration, nonsurvivable physical trauma, and pre-existing medical or neuropsychiatric conditions that could interfere with outcome assessments.

Participants or their legal representatives gave written informed consent. The Galveston Orientation and Amnesia Test (GOAT) was administered to determine ability to consent. For those without a passing score, a legally authorized representative gave initial consent, and competency screening was repeated at all follow-up visits. Race/ethnicity (with options defined by the investigators) were collected to assess for racial/ethnic disparities in outcome that have been reported in prior studies.^17–19^ Institutional review boards of participating centers approved all study protocols.

TRACK-TBI included 2552 adult TBI participants (≥17 years of age), of whom 1078 had the initial clinical head CT, a 2-week standardized-protocol brain MRI, and the GCS score at ED admission available for review (**Figure 1**). Of these 1078 participants, 988 had an ED admission GCS score of 13- 15; and of these, 929 participants had at least one GOSE-TBI outcome score during a 1-year followup period. GOSE-TBI outcome scores were available for 922 participants at 2 weeks (93.3%), 838 at 3 months (84.8%), 804 at 6 months (81.4%), and 754 (76.3%) at 12 months post-injury.

**Figure 1.**
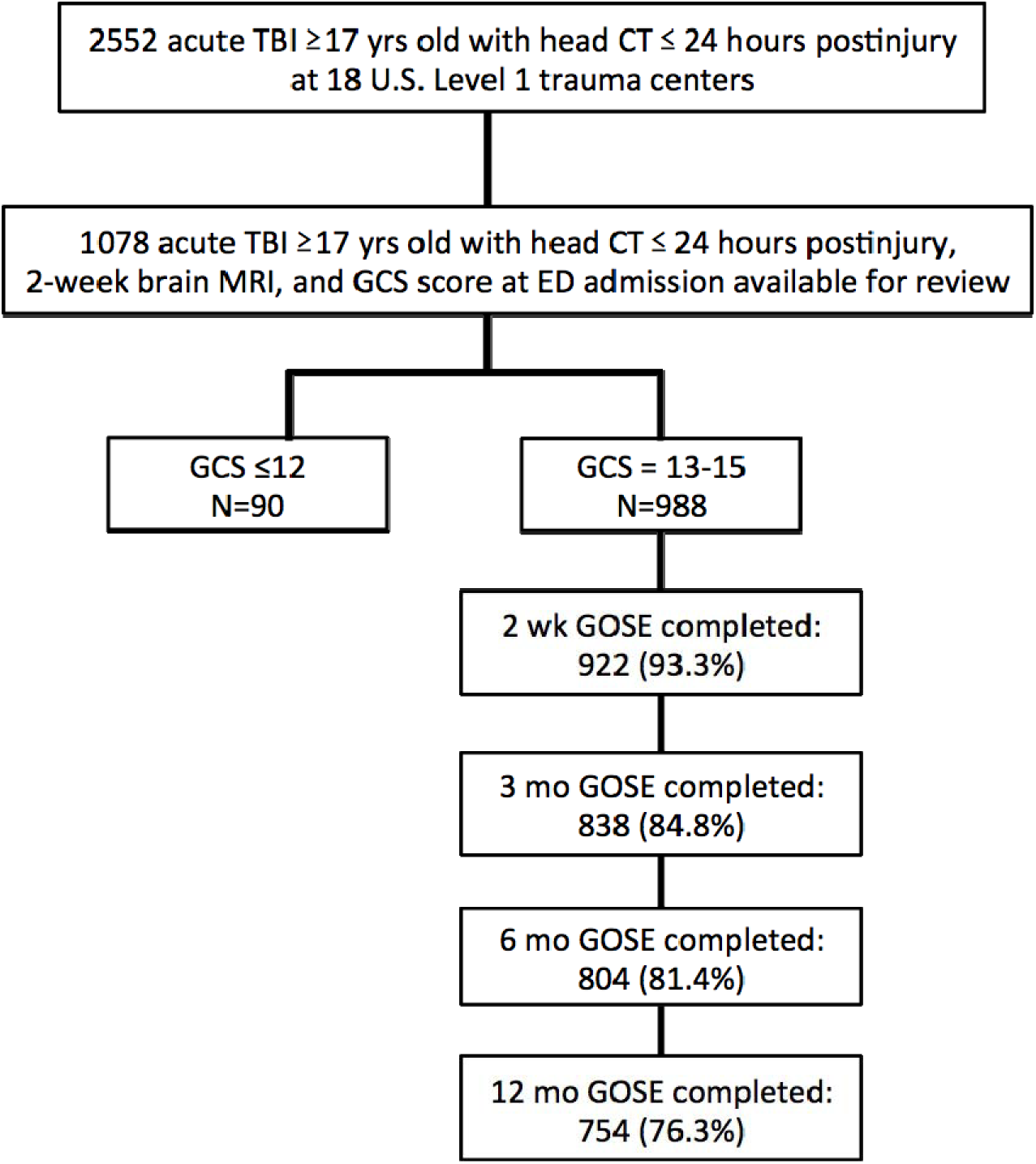
**Recruitment and retention flowchart**

### CT and MR imaging and evaluation of TBI Neuroimaging Common Data Elements

Participants’ initial head CT exams after injury were deidentified, uploaded to a central repository, and evaluated by board-certified neuroradiologists using the National Institute for Neurologic Disorders and Stroke (NINDS) TBI Neuroimaging Common Data Elements (CDE).^20^ Brain MRI exams performed at 2 weeks post-injury were also uploaded to the central repository. Three board- certified neuroradiologists reviewed 517 MRI exams, and all discrepancies were resolved by consensus review by all 3 neuroradiologists.^21^ Two of the neuroradiologists reviewed the remaining 561 MRI exams, and all exams with discrepancies between the two readers regarding the presence or absence of a CDE were resolved through consensus review with the third neuroradiologist. “Traumatic microbleed” (TMB) was used to describe small subcortical or deeper hemorrhages on CT or MRI. TMBs on CT or MRI have been described in the NIH-NINDS TBI Imaging CDEs^20^ to be markers of traumatic (TAI) and diffuse axonal injury (DAI), with DAI defined as 4 or more lesions in at least 2 distinct regions of the brain. ^20^ All neuroradiologist readers were blinded to clinical information except age and sex.

### Outcome measure

The GOSE is the most widely used measure of global functional outcome after TBI.^22–24^ In TRACK-TBI, the primary outcome measure was the GOSE-TBI,^25^ which consists of the GOSE administered with the intent of specifically capturing disability related to the TBI (i.e., excluding any disability attributable to co-occurring polytrauma such as orthopedic injuries).

### Statistical analysis

Descriptive summaries of demographic, clinical, and CT and MRI characteristics of the analysis cohort were provided. Between-group comparisons used Wilcoxon Rank Sum test for continuous variables and Fisher’s exact test for categorical variables. We used hierarchical cluster analysis (HCA) to derive imaging “phenotypes” (clusters of subtypes of intracranial injury) to mitigate collinearity across intracranial lesions seen on both CT and MRI in the same participant.

Generalized Estimating Equations (GEE) were used to study the association of CT and MRI clusters with the GOSE-TBI outcomes at 2 weeks, 3, 6, and 12 months post-injury.^26^ An unstructured working correlation matrix was used. We compared the marginal pseudo-R^2^ statistic for models to assess the contribution of CT or MRI variables. All analyses were performed using R version 4.1.2 (R Foundation for Statistical Computing), using a threshold for statistical significance of p < 0.05, 2- tailed.

## RESULTS

### Demographic and baseline clinical characteristics

#### CT and MR neuroimaging findings

In this cohort study of 929 mTBI patients, traumatic microbleeds were common; ≥4 TMBs in at least 2 distinct regions of the brain, consistent with the NIH-NINDS definition of DAI, were observed in 11.3% of participants on brain MRI at 2 weeks post-injury. **Figure 2** shows results of HCA performed on CT and MR imaging CDEs in the cohort. TMBs on both CT and MRI were more strongly associated with intraventricular hemorrhage (IVH) on CT than with other neuroimaging findings including brain contusion, subarachnoid hemorrhage (SAH), and subdural (SDH) and epidural hematoma (EDH). Brain contusions, SDH, and SAH were more strongly associated with each other than with other findings.

**Figure 2.**
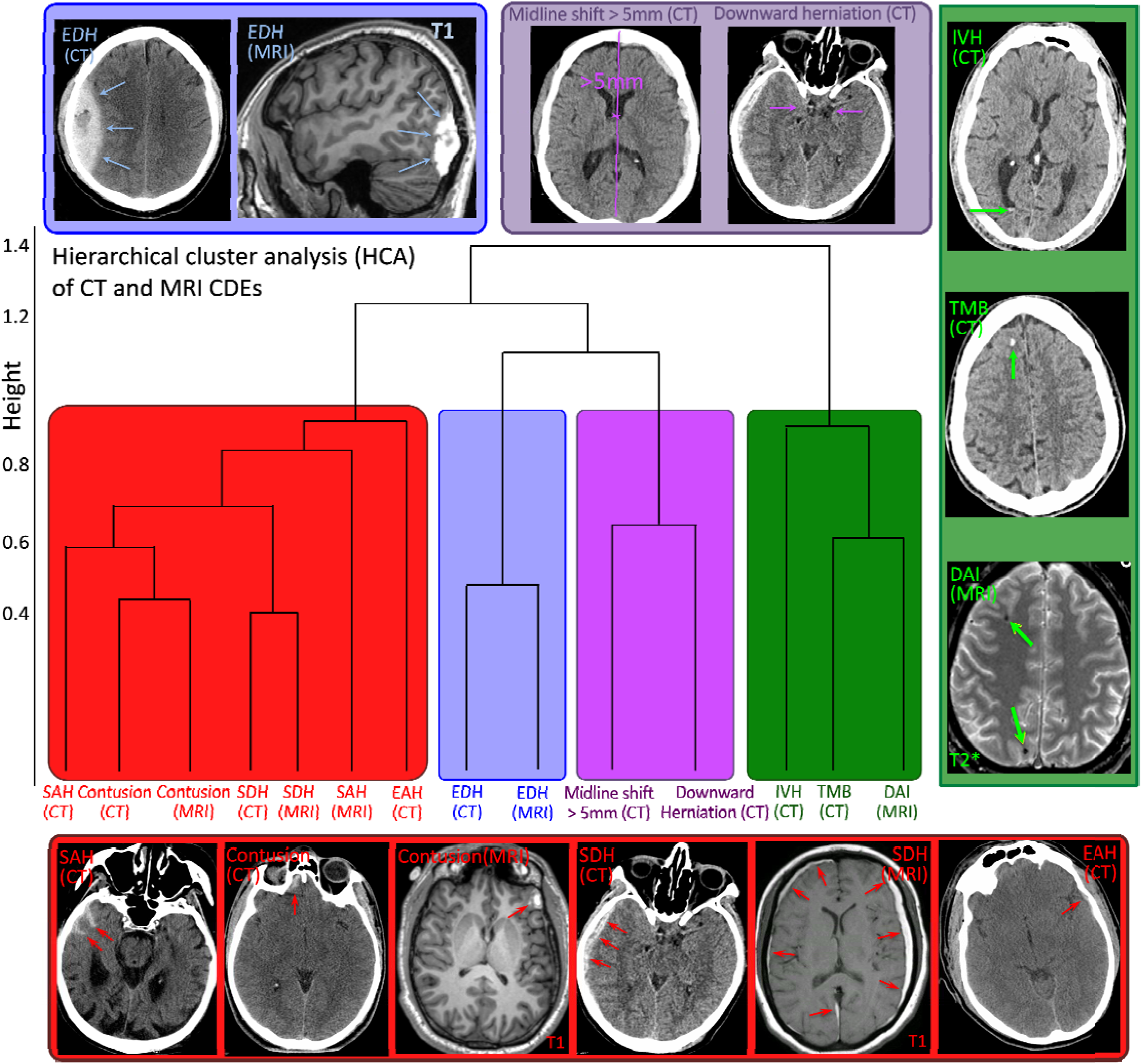
Hierarchical cluster analysis (HCA) results on CT and MRI pathoanatomic imaging Common Data Elements. (**CDEs) (N=929).** Dendrogram demonstrates that CT and MRI Common Data Elements (CD s) are correlated with one another, and based on the tendency for certain CDEs to co-occur, can be viewed as 4 different clusters. *Red cluster:* Brain contusion, subdural hematoma (SDH), or subarachnoid hemorrhage (SAH) on either CT or MRI; and extraaxial hematoma (EAH) on CT. Blue cluster: epidural hematoma (EDH) on C or MRI. *Purple cluster:* Midline shift or downward cerebral herniation on CT. *Green cluster:* any traumatic microbleed (TMB) on CT or diffuse axonal injury (DAI; 4 or more traumatic microbleeds (TMBs)) on MRI, and intraventricular hemorrhage (IVH) on CT. Images along the periphery of the dendrogram show specific examples of the CT and MRI CDEs that appear in the dendrogram.

Finally, imaging features of intracranial mass effect (midline shift and downward herniation) were strongly associated with each other, and weakly associated with the other types of injuries. **Figure 2** also shows typical examples of the CT and MRI appearance of the CDEs in the dendrogram.

From the dendrogram and clinical experience, we defined 4 clusters: 1) contusion on CT or MRI, SAH, SDH and extraaxial hematoma (EAH) on CT, 2) EDH on CT, 3) TMB on CT or MRI, and IVH on CT, and 4) midline shift >5 mm or partial of the basal cisterns (i.e., downward cerebral herniation) on CT. No patient in this mTBI cohort had complete effacement of the basal cisterns. Although SDH and EDH on MRI were strongly associated with their respective counterparts on CT, these were not included in the clusters, as such collections would have resolved in some patients but not in others at 2 weeks post-injury, potentially introducing unwanted confounding effects such as coagulopathy, size of the SDH, and size of the EDH; or random effects such as precise timing of the brain MRI which varied from 11 to 17 days post-injury.

### Association of demographics/baseline clinical features and CT/MRI phenotypes with GOSE-TBI at 2 weeks and 3, 6 and 12 months post-injury

GEE models assessed the association of CT/MRI phenotypes and demographic and baseline clinical features with incomplete recovery (GOSE-TBI <8 vs. =8; **Table 2** and **Figure 3, left panel**) and unfavorable outcome (GOSE-TBI <5 vs. ≥5; **Table 3** and **Figure 3, right panel**) at 2 weeks and 3, 6, and 12 months post-injury.

**Figure 3.**
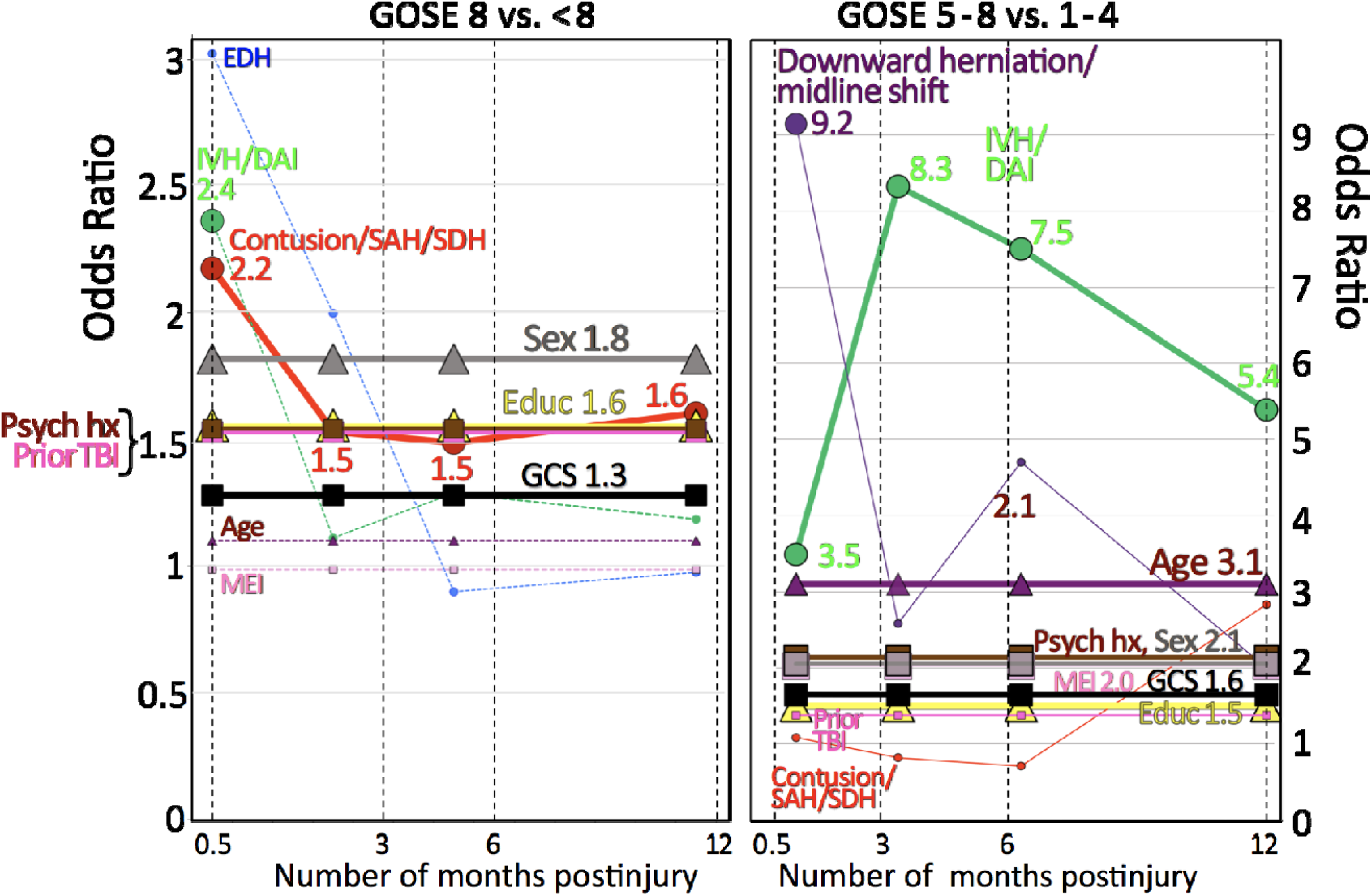
Odds ratios of predictors of incomplete recovery (GOSE-TBI <8) and unfavorable outcome (GOSE-TBI <5) at 2 weeks and 3, 6 and 12 months post-injury for participants with ED admission GCS scores of 13-15. Each number next to a large marker (triangle, square, or circle) denotes a statistically significant odds ratio (from **Tables 2 and 3**) for participants with a specific demographic, baseline clinical, or CT/MRI feature to experience incomplete recovery (left panel: GOSE-TBI <8 vs. =8) or unfavorable outcome (right panel: GOSE-TBI <5 vs. ≥5) at a given time point. Large markers (triangle, square, or circle) are used to emphasize only statistically significant odds ratios (p<0.05) from **Tables 2 and 3**. Small markers (also triangle, square, or circle) denote odds ratios from **Tables 2 and 3** with p>0.05. Triangles are used to represent demographic features (a e, education, sex), squares for baseline clinical features (admission GCS score, history of prior TBI, history of neuropsychiatric disorder, concurrent major extracranial injury), and circles for CT/MRI features (EDH, SDH, contusion/SAH/SDH, IVH/DAI, downward herniation/midline shift).

**Table 1.**
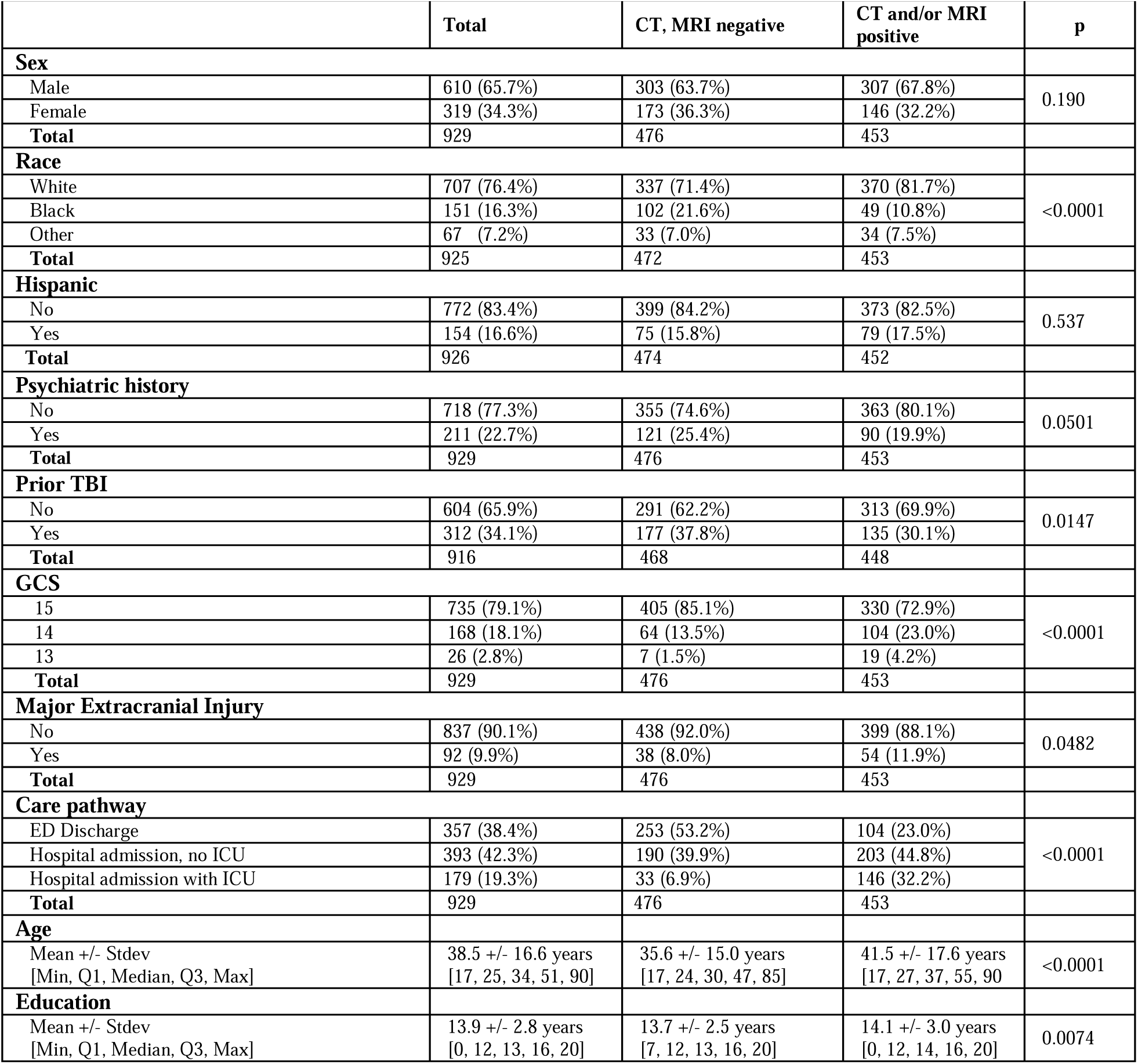
presents demographic and baseline clinical characteristics of the cohort evaluated in the present study: TRACK-TBI participants aged ≥17 years with ED admission GCS score of 13-15, with initial head CT and 2-week brain MRI available for review, and with at least one GOSE-TBI score during the 1-year follow-up period (N=929). Most participants were male (65.7%). The most common clinical care pathway was hospital admission without ICU stay (42.3%), followed by discharge from the ED (38.4%) and hospital admission with ICU stay (19.3%). Participants with evidence of acute traumatic intracranial injury on CT or MRI were older (p<0.001), had more years of education (p=0.007), and were more likely to be White (p<0.001), have a prior TBI history (p=0.01), and be treated under the ICU care pathway (p<0.001).

|  | **Total** | **CT, MRI negative** | **CT and/or MRI positive** | **p** |
|---|---|---|---|---|
| **Sex** |  |  |  |  |
| Male | 610 (65.7%) | 303 (63.7%) | 307 (67.8%) | 0.190 |
| Female | 319 (34.3%) | 173 (36.3%) | 146 (32.2%) |  |
| **Total** | 929 | 476 | 453 |  |
| **Race** |  |  |  |  |
| White | 707 (76.4%) | 337 (71.4%) | 370 (81.7%) | <0.0001 |
| Black | 151 (16.3%) | 102 (21.6%) | 49 (10.8%) |  |
| Other | 67 (7.2%) | 33 (7.0%) | 34 (7.5%) |  |
| **Total** | 925 | 472 | 453 |  |
| **Hispanic** |  |  |  |  |
| No | 772 (83.4%) | 399 (84.2%) | 373 (82.5%) | 0.537 |
| Yes | 154 (16.6%) | 75 (15.8%) | 79 (17.5%) |  |
| **Total** | 926 | 474 | 452 |  |
| **Psychiatric history** |  |  |  |  |
| No | 718 (77.3%) | 355 (74.6%) | 363 (80.1%) | 0.0501 |
| Yes | 211 (22.7%) | 121 (25.4%) | 90 (19.9%) |  |
| **Total** | 929 | 476 | 453 |  |
| **Prior TBI** |  |  |  |  |
| No | 604 (65.9%) | 291 (62.2%) | 313 (69.9%) | 0.0147 |
| Yes | 312 (34.1%) | 177 (37.8%) | 135 (30.1%) |  |
| **Total** | 916 | 468 | 448 |  |
| **GCS** |  |  |  |  |
| 15 | 735 (79.1%) | 405 (85.1%) | 330 (72.9%) | <0.0001 |
| 14 | 168 (18.1%) | 64 (13.5%) | 104 (23.0%) |  |
| 13 | 26 (2.8%) | 7 (1.5%) | 19 (4.2%) |  |
| **Total** | 929 | 476 | 453 |  |
| **Major Extracranial Injury** |  |  |  |  |
| No | 837 (90.1%) | 438 (92.0%) | 399 (88.1%) | 0.0482 |
| Yes | 92 (9.9%) | 38 (8.0%) | 54 (11.9%) |  |
| **Total** | 929 | 476 | 453 |  |
| **Care pathway** |  |  |  |  |
| ED Discharge | 357 (38.4%) | 253 (53.2%) | 104 (23.0%) | <0.0001 |
| Hospital admission, no ICU | 393 (42.3%) | 190 (39.9%) | 203 (44.8%) |  |
| Hospital admission with ICU | 179 (19.3%) | 33 (6.9%) | 146 (32.2%) |  |
| **Total** | 929 | 476 | 453 |  |
| **Age** |  |  |  |  |
| Mean +/- Stdev<br>[Min, Q1, Median, Q3, Max] | 38.5 +/- 16.6 years<br>[17, 25, 34, 51, 90] | 35.6 +/- 15.0 years<br>[17, 24, 30, 47, 85] | 41.5 +/- 17.6 years<br>[17, 27, 37, 55, 90 | <0.0001 |
| **Education** |  |  |  |  |
| Mean +/- Stdev<br>[Min, Q1, Median, Q3, Max] | 13.9 +/- 2.8 years<br>[0, 12, 13, 16, 20] | 13.7 +/- 2.5 years<br>[7, 12, 13, 16, 20] | 14.1 +/- 3.0 years<br>[0, 12, 14, 16, 20] | 0.0074 |

**Table 2.** Association of demographic, baseline clinical, and CT/MRI phenotypes with incomplete recovery (GOSE-TBI <8 vs. =8) at 2 weeks and 3, 6 and 12 months post-injury based on generalized estimating equations (GEE)

| **Predictors** | **post-injury time point** | **OR** | **95% CI** | **p-value** |
|---|---|---|---|---|
| **CT/MRI Phenotypes** |  |  |  |  |
| Contusion/SAH/SDH | Week 2 | 2.176 | (1.441, 3.286) | <0.001*** |
|  | Month 3 | 1.532 | (1.097, 2.141) | 0.012* |
|  | Month 6 | 1.487 | (1.062, 2.081) | 0.021* |
|  | Month 12 | 1.603 | (1.149, 2.236) | 0.005** |
| EDH | Week 2 | 3.022 | (0.897, 10.187) | 0.074 |
|  | Month 3 | 1.997 | (0.934, 4.270) | 0.074 |
|  | Month 6 | 0.900 | (0.455, 1.779) | 0.761 |
|  | Month 12 | 0.975 | (0.489, 1.944) | 0.944 |
| IVH/TMB/DAI | Week 2 | 2.365 | (1.274, 4.388) | 0.006** |
|  | Month 3 | 1.112 | (0.720, 1.718) | 0.633 |
|  | Month 6 | 1.289 | (0.852, 1.949) | 0.23 |
|  | Month 12 | 1.185 | (0.782, 1.796) | 0.422 |
| **Demographics** |  |  |  |  |
| Age (50 vs 25 yrs) |  | 1.101 | (0.928, 1.306) | 0.27 |
| Years of Education (12 vs 16 yrs) |  | 1.555 | (1.321, 1.832) | <0.001*** |
| Sex (Female vs Male) |  | 1.819 | (1.442, 2.295) | <0.001*** |
| **Baseline Clinical Characteristics** |  |  |  |  |
| Neuropsychiatric history (Yes vs No) |  | 1.542 | (1.185, 2.007) | 0.001** |
| Prior TBI (Yes vs No) |  | 1.530 | (1.220, 1.920) | <0.001*** |
| GCS (13 vs 15) |  | 1.281 | (1.033, 1.588) | 0.024* |
| Major Extracranial Injury (Yes vs No) |  | 0.987 | (0.683, 1.427) | 0.945 |

**Table 3.** Association of demographic, baseline clinical, and CT/MRI phenotypes with unfavorable outcome (GOSE-TBI <5 vs. ≥5) at 2 weeks and 3, 6 and 12 months post-injury

| **Predictors** | **post-injury time point** | **OR** | **95% CI** | **p-value** |
|---|---|---|---|---|
| **CT/MRI Phenotypes** |  |  |  |  |
| Contusion/SAH/SDH | Week 2 | 1.085 | (0.579, 2.033) | 0.799 |
|  | Month 3 | 0.816 | (0.226, 2.943) | 0.756 |
|  | Month 6 | 0.708 | (0.160, 3.138) | 0.649 |
|  | Month 12 | 2.827 | (0.717, 11.148) | 0.138 |
| IVH/TMB/DAI | Week 2 | 3.490 | (1.703, 7.153) | <0.001*** |
|  | Month 3 | 8.325 | (2.376, 29.171) | <0.001*** |
|  | Month 6 | 7.500 | (1.826, 30.801) | 0.005** |
|  | Month 12 | 5.392 | (1.612, 18.033) | 0.006** |
| Midline shift/Downward cerebral herniation | Week 2 | 9.150 | (2.938, 28.498) | <0.001*** |
|  | Month 3 | 2.580 | (0.397, 16.769) | 0.321 |
|  | Month 6 | 4.703 | (0.877, 25.223) | 0.071 |
|  | Month 12 | 1.998 | (0.388, 10.293) | 0.408 |
| **Demographics** |  |  |  |  |
| Age (50 vs 25 yrs) |  | 3.098 | (2.105, 4.559) | <0.001*** |
| Years of Education (12 vs 16 yrs) |  | 1.500 | (1.038, 2.166) | 0.031* |
| Sex (Female vs Male) |  | 2.048 | (1.103, 3.802) | 0.023* |
| **Baseline Clinical Characteristics** |  |  |  |  |
| Neuropsychiatric history (Yes vs No) |  | 2.136 | (1.138, 4.009) | 0.018* |
| Prior TBI (Yes vs No) |  | 1.377 | (0.764, 2.483) | 0.288 |
| GCS (13 vs 15) |  | 1.644 | (1.046, 2.582) | 0.031* |
| Major Extracranial Injury (Yes vs No) |  | 2.018 | (1.039, 3.919) | 0.038* |

The addition of imaging features improved GEE model accuracy for both incomplete recovery and unfavorable outcome out to 1 year post-injury. As shown in **Table 2**, the marginal R^2^ of the GEE model for incomplete recovery (GOSE-TBI <8) improved from 10.2% without CT or MRI features to 11.9% with these features. For unfavorable outcome (GOSE-TBI <5), the marginal R^2^ of the GEE model improved from 7.0% without CT or MRI features to 13.5% with these features. Addition of MRI findings to only CT findings in the GOSE-TBI <5 model resulted in a small marginal R^2^ increase, from 12.0% (with only CT features) to 13.5% (with both CT and MRI features). Addition of MRI findings to only CT findings in the GOSE-TBI <8 model did not result in a change in the marginal R^2^ (11.9% with only CT features and 11.9% with both CT and MRI features).

For prediction of incomplete recovery (GOSE-TBI <8 vs. =8; **Table 2** and **Figure 3, left panel**), contusion/SAH/SDH were of greater importance than any single socio-demographic or baseline clinical characteristic early on (at 2 weeks), and of comparable importance at later time points (3, 6, and 12 months). Specifically, odds ratios for contusion/SAH/SDH were in the 1.5 to 2.2 range over the followup period, while odds ratios for admission GCS, sex, years of education, history of neuropsychiatric disorder, history of prior TBI were in the 1.3 to 1.8 range. Admission GCS was significantly associated with GOSE-TBI, with an odds ratio of 1.3 comparing GCS=13 vs GCS=15.

For prediction of greater degrees of unfavorable outcome (GOSE-TBI <5 vs. GOSE-TBI ≥5; **Table 3** and **Figure 3, right panel**), imaging features associated with rotational acceleration (IVH/TMBs) greatly increased in importance, with odds ratios of 3.5 to 8.3, surpassing all socio- demographic and baseline clinical predictors throughout the 12 month followup period. Although imaging features associated with intracranial mass effect fell short of statistical significance except at the 2-week time point, this is likely due to the relatively low prevalence of these imaging features and GOSE-TBI scores <5 in the GCS 13-15 subgroup, rather than decreased importance: the odds ratios for these features, though not significant at p<0.05, are relatively large.

### Generalized estimating equation (GEE) model was used to study the association of demographics, clinical, and CT/MRI predictors with incomplete recovery (GOSE-TBI <8 vs. =8) at 2 weeks, 3, 6, and 12 months post- injury

The model included GOSE-TBI (<8 vs. =8) at each follow-up as the outcome; predictors included demographics, baseline clinical characteristics, CT/MRI phenotypes, timepoints, and interaction between CT/MRI phenotypes and timepoints. An unstructured working correlation matrix was used. Note for the continuous predictors (age and years of education), we reported OR comparing its third quartile vs. first quartile. The marginal R^2^ of the GEE model was 10.2% without CT or MRI predictors, 11.9% with only CT predictors, and 11.9% with CT/MRI predictors. Midline shift and downward cerebral herniation were not included in this model for GOSE =8 vs. <8 because nearly all participants with midline shift and/or downward cerebral herniation had GOSE<8 at 2 weeks and 3 months.

Generalized estimating equation (GEE) model was used to study the association of demographics, clinical, and CT/MRI predictors with unfavorable outcome (GOSE-TBI <5 vs. ≥5) at 2 weeks, 3, 6, and 12 months post-injury. The model included GOSE-TBI (<5 vs. ≥5) at each follow-up as the outcome; predictors included demographics, baseline clinical characteristics, CT/MRI phenotypes, timepoints, and interaction between CT/MRI phenotypes and timepoints. An unstructured working correlation matrix was used. Note for the continuous predictors (age and years of education), we reported OR comparing its third quartile vs. first quartile. The marginal R^2^ of the GEE model was 7.0% without CT or MRI predictors, 12.0% with only CT predictors, and 13.5% with CT/MRI predictors. EDH was not included in this model for GOSE <5 vs. 5 as there were no subjects with EDH and GOSE <5 at 3, 6 or 12 months.

## DISCUSSION

In this study, we sought to determine the long-term prognostic significance of TMBs in mild TBI. We found that this varied widely at different points across the injury spectrum. For prediction of incomplete recovery (GOSE-TBI <8 vs. =8), ≥4 TMBs on brain MRI, or any TMB or IVH on head CT, were significant only at the 2-week post-injury time point, but not at the 3, 6, or 12 month post-injury time points. In contrast, for prediction of greater degrees of unfavorable outcome (GOSE-TBI <5 vs.

GOSE-TBI ≥5), TMBs greatly increased in importance. Specifically, the presence of ≥4 TMBs on brain MRI, or any TMB or IVH on head CT, were strongly associated with unfavorable outcome (GOSE-TBI <5) out to 12 months after mTBI, with odds ratios (OR) of 8.3 [2.4-29.2], 7.5 [1.8-30.8], and 5.4 [1.6- 18.0] for GOSE-TBI <5 vs. ≥5 at 3, 6, and 12 months, respectively. These far exceeded the ORs for admission GCS score (13 vs15), age, sex, history of neuropsychiatric disorder, history of prior TBI, and concurrent major extracranial injury.

Based on HCA, we found that IVH and TMBs, whether identified on CT or MRI, co-occurred much more frequently with each other than with other intracranial imaging abnormalities. Brain contusions, SDH, and SAH co-occurred more frequently with each other than with IVH or TMBs.

Finally, imaging features of intracranial mass effect (midline shift and downward herniation) associated strongly with each other, and much less strongly with the other types of injuries.

Griffin et al.^10^ demonstrated that TMBs, though more prevalent in moderate to severe TBI (GCS ≤12), are common across all GCS scores. They used binary logistic regression to show TMBs are associated with worse outcome (GOSE-TBI ≤6) at intermediate-term follow-up (1 to 3 months), even after adjusting for GCS score (3-15) and positive head CT. Strengths of the current study include a larger sample size (N=929 vs. N=250) and longer follow-up period (1 year vs. 1 to 3 months post- injury), providing a picture of longer-term outcome. Other differences are that the present study concerns admission GCS scores of 13-15 only, limiting the potential for confounding or otherwise unaccounted-for factors such as surgical management. In addition, the distribution of GCS scores in the current study was skewed higher than that of the mTBI subgroup in Griffin et al.,^10^ toward “milder” TBI: the significantly higher admission GCS scores in the current study (79% vs. 70% with GCS 15) more closely match the distribution in acute mTBI in U.S. hospitals.^27^ Finally, statistical analysis in the present study differed: generalized estimating equations were used to take advantage of expected intra- participant correlations in repeated GOSE-TBI measures across time; and we used two cutpoints, at GOSE-TBI <5 and GOSE-TBI <8, which was essential in order to uncover the varying clinical significance of CT and MR pathological findings for functional outcome at different points along the injury severity spectrum.

Otherwise, the present study is congruent with Griffin et al.^10^ regarding the prevalence of microbleeds in acute mTBI. Prevalence of ≥1TMB in the present study, 29% in GCS 13-15 participants, was very similar to that in Griffin et al. (27% for their mild TBI subgroup).^10^ Interrater reliability of neuroradiologists for TMBs was also similar between the two studies.^10,21^

TMBs have been associated with rotational acceleration in animal models.^28^ In our study, IVH was strongly associated with TMBs on both CT and MRI in HCA, and is thus presumed to also be associated with rotational acceleration resulting in hemorrhage from traumatic injury to subependymal or periventricular blood vessels. Based on the very deep location of these blood vessels associated with the ventricular system, IVH presumably requires higher rotational acceleration in order to achieve the same linear velocity in deeper structures as at the peripheral location of the more common subcortical hemorrhages in lobar white matter.

A number of other studies have focused on the clinical significance of TMBs on MRI specifically in mTBI patients (GCS 13-15). Although there has been some evidence of an association between TMBs on MRI and higher symptom severity at 1 year in mTBI,^11,13^ most studies have not demonstrated an association. Einarsen et al.^9^ found no statistically significant association of TMBs with postconcussive symptoms in mTBI in 183 patients at 3 months or in 167 patients at 1 year. Similarly, Huovinen et al.^12^ studied 113 mTBI patients with MRI at 3-17 days post-injury, and found no difference in GOSE or return-to-work at 3 months. Van der Horn et al.^6^ showed that TMBs, while common even in CT-negative mTBI (referred to as “uncomplicated” mTBI^29^ in that study), had no association with outcome in 54 CT-negative patients, even at 2 weeks post-injury. Although the results presented here may appear to conflict with these studies, a reasonable explanation is that the prior studies did not have sufficient power to detect a significant association with GOSE at p<0.05. Einarsen et al.^9^ performed power calculations that suggested at least 300 or more mTBI patients would have been required to demonstrate a statistically significant effect at p<0.05, based on the prevalence of postconcussive symptoms in their study sample. This greatly exceeds the numbers of patients evaluated in any of these studies. Furthermore, the mTBI patients in Huovinen et al.^12^ sustained significantly milder injuries than the TRACK-TBI mTBI subgroup: 56% attained a GOSE score of 8 by 3 months post-injury, compared to only 40% in TRACK-TBI; and only 34% of mTBI patients had any traumatic intracranial findings on brain MRI, compared to 43% in TRACK-TBI patients with GCS 13-15.

Finally, we also sought in this study to determine the additive prognostic value of early brain MRI when added to head CT findings. A prior study claimed that MRI improves prediction of 3-month GOSE in mTBI, based on comparison of OLR models of 3-month outcome without or with MRI predictors (added to a baseline model containing only sociodemographic, clinical, and CT predictors).^15^ The present study goes further by analyzing longer-term outcome (6 and 12 months postinjury).

Addition of MRI findings to only CT findings in the GEE model for GOSE-TBI <5 out to 1 year resulted in an R^2^ increase, from 12.0% (with only CT features) to 13.5% (with both CT and MRI features) (**Table 3**). The model for incomplete recovery (GOSE-TBI <8 vs. =8) out to 1 year demonstrated no R^2^ change when MRI findings were added to CT findings (**Table 2**). Although the R^2^ increase from 12.0% to 13.5% for the GOSE-TBI <5 model is relatively modest, the odds ratios (OR) for the association of ≥4 TMBs on brain MRI, or any TMB or IVH on head CT, with GOSE-TBI <5 at 3, 6 and 12 months postinjury were strikingly large (8.3 [2.4-29.2], 7.5 [1.8-30.8], and 5.4 [1.6-18.0]), and far exceeded ORs for other predictors, including admission GCS score (13 vs 15), age, sex, history of neuropsychiatric disorder, history of prior TBI, and concurrent major extracranial injury. This shows that these specific imaging findings are highly significant, accounting for a disproportionately large number of mTBI patients with long-term unfavorable outcome (GOSE-TBI <5).

## CONCLUSION

Traumatic microbleeds (TMB) are common in mTBI. We showed that rotational acceleration injuries in “mild” TBI patients, while less common than in “moderate to severe” TBI, can have profound implications for outcome. The presence of ≥4 TMBs and/or IVH was strongly associated with unfavorable outcome (GOSE-TBI <5 vs. ≥5), with odds ratios ranging from 5.4 to 8.3 at 3, 6, and 12 months post-injury, surpassing the importance of all demographic and baseline clinical predictors throughout the 1-year follow-up period. The odds ratios decrease over time, and this may be at least partly due to the longer time period for unaccounted-for factors (e.g., access to rehabilitation resources, social support, brain reserve, cognitive reserve, and other contributors to individual resilience) to influence outcome as time progresses. However, they remain high and statistically significant at 12 months postinjury. This highlights the potential for focusing more supportive resources for mTBI patients with TMBs to address their strong tendency toward the poorest long-term outcomes.

### The TRACK-TBI Investigators

Ann-Christine Duhaime, MD, Mass General Brigham for Children; Brandon Foreman, MD, University of Cincinnati; Shankar Gopinath, MD, Baylor College of Medicine; Ramesh Grandhi, MD MS, University of Utah; Ruchira Jha, MD, MSc, Barrow Neurological Institute; C. Dirk Keene, MD PhD, Christopher Madden, MD, UT Southwestern; Alastair Martin, PhD, University of California, San Francisco; Randall Merchant, PhD, Virginia Commonwealth University; Laura B. Ngwenya, MD, PhD, University of Cincinnati; Andrea Schneider, MD PhD, University of Pennsylvania; David Schnyer, PhD, UT Austin; Ross Zafonte, DO, Harvard Medical School

## Data Availability

All data produced in the present study are available upon reasonable request to the authors

## SUPPLEMENTARY MATERIAL

| **Transforming Clinical Knowledge in Traumatic Brain Injury (TRACK-TBI) sites** |
|---|
| Ben Taub General Hospital in Houston, TX |
| Massachusetts General Hospital in Boston, MA |
| Zuckerberg San Francisco General in San Francisco, CA |
| University of Cincinnati Medical Center in Cincinnati, OH |
| R. Adams Cowley Shock Trauma Center in Baltimore, MD |
| Ryder Trauma Center in Miami, FL |
| University of Pittsburgh Medical Center in Pittsburgh, PA |
| Seton Medical Center in Austin, TX |
| Parkland Memorial Hospital in Dallas, TX |
| Harborview Medical Center in Seattle, WA |
| Virginia Commonwealth University Medical Center in Richmond, VA |
| University of Pennsylvania Health System in Philadelphia, PA |
| Emory Healthcare in Atlanta, GA |
| Medical College of Wisconsin in Milwaukee, WI |
| University of Utah Health in Salt Lake City, UT |
| Indiana University Health in Indianapolis, IN |
| Hennepin County Medical Center in Minneapolis, MN |
| Denver Health in Denver, CO |

## REFERENCES

1. Hughes DG, Jackson A, Mason DL, Berry E, Hollis S, Yates DW. Abnormalities on magnetic resonance imaging seen acutely following mild traumatic brain injury: correlation with neuropsychological tests and delayed recovery. Neuroradiology. 2004;46:550–558.

2. Lee H, Wintermark M, Gean AD, Ghajar J, Manley GT, Mukherjee P. Focal lesions in acute mild traumatic brain injury and neurocognitive outcome: CT versus 3T MRI. J Neurotrauma. 2008;25(9):1049–1056.

3. Scheid R, Preul C, Gruber O, Wiggins C, von Cramon DY. Diffuse axonal injury associated with chronic traumatic brain injury: Evidence from T2*-weighted gradient-echo imaging at 3 T. AJNR Am J Neuroradiol. 2003;24:1049–1056.

4. de Haan S. dGJC, Jacobs B., and van der Naalt J. The association between microhaemorrhages and post-traumatic functional outcome in the chronic phase after mild traumatic brain injury. Neuroradiology. 2017;59:963–969.

5. Iverson GL, Lange, R.T., Wäljas, M., Liimatainen, S., Dastidar, P., Hartikainen, K.M., Soimakallio, S. and Öhman, J. Outcome from complicated versus uncomplicated mild traumatic brain injury. Rehabilitation research and practice. 2012.

6. Van der Horn HJ, de Haan, S., Spikman, J.M., de Groot, J.C. and van der Naalt, J. Clinical relevance of microhemorrhagic lesions in subacute mild traumatic brain injury. Brain Imaging and Behavior. 2018;12(3):912–916.

7. Wäljas M, Iverson GL, Lange RT, et al. A prospective biopsychosocial study of the persistent post-concussion symptoms following mild traumatic brain injury. Journal of neurotrauma. 2015;32(8):534–547.

8. Panenka WJ, Lange, R.T., Bouix, S., Shewchuk, J.R., Heran, M.K., Brubacher, J.R., Eckbo, R., Shenton, M.E. and Iverson, G.L. Neuropsychological outcome and diffusion tensor imaging in complicated versus uncomplicated mild traumatic brain injury. PLoS One. 2015;10(4):e0122746.

9. Einarsen CE MK, Håberg AK, Eikenes L, Kvistad KA, Xu J. Patients with mild traumatic brain injury recruited from both hospital and primary care settings: a controlled longitudinal magnetic resonance imaging study. J Neurotrauma. 2019;36:3172–3182.

10. Griffin AD, Turtzo, L.C., Parikh, G.Y., Tolpygo, A., Lodato, Z., Moses, A.D., Nair, G., Perl, D.P., Edwards, N.A., Dardzinski, B.J. and Armstrong, R.C. Traumatic microbleeds suggest vascular injury and predict disability in traumatic brain injury. Brain. 2019;142(11):3550–3564.

11. Huang YL KY, Tseng YC, Chen DY, Chiu WT, Chen CJ. Susceptibility-weighted MRI in mild traumatic brain injury. Neurology. 2015;84:580–585.

12. Huovinen A, Marinkovic, I., Isokuortti, H., Korvenoja, A., Mäki, K., Nybo, T., Raj, R. and Melkas, S. Traumatic microbleeds in mild traumatic brain injury are not associated with delayed return to work or persisting post-concussion symptoms. J Neurotrauma. 2021;38(17):2400–2406.

13. Studerus-Germann AM GO, Bontempi P, Thiran JP, Daducci A, Romascano D. Central nervous system microbleeds in the acute phase are associated with structural integrity by DTI one year after mild traumatic brain injury: a longitudinal study. Neurol Neurochir Polska. 2018;52(710-719).

14. Yuh EL, Cooper SR, Mukherjee P, et al. Diffusion tensor imaging for outcome prediction in mild traumatic brain injury. J Neurotrauma. 2014;31:1457–1477.

15. Yuh EL, Mukherjee P, Lingsma HF, et al. Magnetic resonance imaging improves 3-month outcome prediction in mild traumatic brain injury. Annals of neurology. 2013;73(2):224–235.

16. Jagoda AS, Bazarian JJ, Bruns JJ, et al. Clinical policy: Neuroimaging and decisionmaking in adult mild traumatic brain injury in the acute setting. Ann Emerg Med. 2008;52(6):714–748.

17. Meagher AD, Beadles CA, Doorey J, Charles AG. Racial and ethnic disparities in discharge to rehabilitation following traumatic brain injury. Journal of neurosurgery. 2015;122(3):595–601.

18. Shafi S, de la Plata CM, Diaz-Arrastia R, et al. Racial disparities in long-term functional outcome after traumatic brain injury. Journal of Trauma and Acute Care Surgery. 2007;63(6):1263–1270.

19. Staudenmayer KL, Diaz-Arrastia R, de Oliveira A, Gentilello LM, Shafi S. Ethnic disparities in long-term functional outcomes after traumatic brain injury. Journal of Trauma and Acute Care Surgery. 2007;63(6):1364–1369.

20. Haacke EM, Duhaime AC, Gean AD, et al. Common data elements in radiologic imaging of traumatic brain injury. J Magn Reson Imaging. 2010;32(3):516–543.

21. Rincon SP, Mukherjee, P., Levin, H.S., Temkin, N.R., Mac Donald, C.L., Krainak, D.M., Sun, X., Jain, S., Taylor, S.R., Markowitz, A.J., Kumar, A., Manley, G.T., Yuh E.L. Interrater reliability of National Institutes of Health traumatic brain injury imaging common data elements for brain magnetic resonance imaging in mild traumatic brain injury. J Neurotrauma. 2021;38(20):2831–2840.

22. Hudak AM, Caesar RR, Frol AB, et al. Functional outcome scales in traumatic brain injury: a comparison of the Glasgow Outcome Scale (Extended) and the Functional Status Examination. J Neurotrauma. 2005;22(11):1319–1326.

23. Levin HS, Boake C, Song J, et al. Validity and sensitivity to change of the Extended Glasgow Outcome Scale in Mild to Moderate Traumatic Brain Injury. J Neurotrauma. 2004;18(6):575–584.

24. Wilson JT, Pettigrew LE, Teasdale GM. Structured interviews for the Glasgow Outcome Scale and the extended Glasgow Outcome Scale: guidelines for their use. J Neurotrauma. 1998;15:573–585.

25. Wilson L, Boase, K., Nelson, L.D., Temkin, N.R., Giacino, J.T., Markowitz, A.J., Maas, A., Menon, D.K., Teasdale, G. and Manley, G.T. A manual for the glasgow outcome scale- extended interview. J Neurotrauma. 2021;38(17):2435–2446.

26. Zheng B. Summarizing the goodness of fit of generalized linear models for longitudinal data. Statistics in medicine. 2000;19(10):1265–1275.

27. Korley FK, Kelen GD, Jones CM, Diaz-Arrastia R. Emergency department evaluation of traumatic brain injury in the United States, 2009–2010. The Journal of head trauma rehabilitation. 2016;31(6):379.

28. Ommaya AK, Gennarelli TA. Cerebral concussion and traumatic unconsciousness. Correlation of experimental and clinical observations of blunt head injuries. Brain. 1974;97(4):633–654.

29. Williams DH, Levin HS, Eisenberg HM. Mild head injury classification. Neurosurgery. 1990;27:422–428.

